# Feasibility of 24-hour physical behavior monitoring with thigh-worn accelerometers after childhood cancer – the Swiss Childhood Cancer Survivor Study (SCCSS) - Activity

**DOI:** 10.1101/2025.10.04.25337334

**Authors:** C Nigg, D Mohr, C Schindera, E Brack, CE Kuehni

## Abstract

**Background:** A healthy lifestyle, including sufficient physical activity, limited sedentary behavior, and adequate sleep (24-hour physical behaviors) can mitigate late effect risks for childhood cancer survivors (CCS). Most research on 24-hour physical behaviors has been questionnaire-based, while device-based assessment is lacking. Here, we assess the feasibility of 24-hour physical behavior monitoring using thigh-worn accelerometers in an age-diverse CCS population.

**Methods:** SCCSS-Activity is nested in the Swiss Childhood Cancer Survivor Study (SCCSS), a nation-wide questionnaire-based cohort study. As part of the 2024/2025 mailing, we asked participants if they are willing to participate in an accelerometer-based sub-study. Participants wore an activPAL4® accelerometer continuously for eight days. We investigated feasibility using the following indicators: recruitment rate, acceptability and suitability of contact procedures and measurement scheduling, accelerometer return and adherence, summary reports, and overall impression. We analyzed quantitative data descriptively and conducted semi-structured interviews with nine participants.

**Results:** Of 367 CCS returning the baseline questionnaire (median: 14 years [IQR: 10-21]; 50% female), 157 (43%) expressed interest in the accelerometer study, 100 were enrolled as of June 2025, and 90 wore and returned the accelerometer, with all except two containing valid data. Participants wore the device for a median of seven days (IQR 7–8). Interview results indicated high acceptability and minimal burden.

**Conclusion:** Twenty-four-hour physical behavior assessment using thigh-worn accelerometers is feasible and well accepted among pediatric, adolescent, and adult CCS. This holds promise for future research to better understand and intervene on physical behaviors and to understand their compositional impact on CCS’ health.

**Highlights:**

- Thigh-worn accelerometry is feasible to assess 24-hour physical behaviors in survivors
- 43% of eligible survivors expressed interest in accelerometer participation
- 98% of participants provided valid accelerometry data
- Mailing-based recruitment and return procedures were effective and acceptable
- Survivors reported low burden and high acceptability of study participation

## Introduction

Five-year childhood cancer survival rates in Switzerland approach 90% (1). Nevertheless, two-thirds of childhood cancer survivors (CCS) experience late effects related to their cancer or its treatment that can impair long-term health and quality of life (2–7). Health-promoting 24-hour physical behaviors, including sufficient physical activity and sleep, and limited sedentary behavior, can mitigate several of those late effects (8), and often occur together with other health-promoting behaviors (e.g., a healthy diet) (9) that also mitigate late effect risk. However, we showed previously that adult CCS engage in less exercise than healthy controls (10), especially if they have physical performance limitations (e.g., musculoskeletal or cardiorespiratory problems) (11), and that adolescent CCS tend to exceed screen time recommendations (12). Findings from other studies confirm that CCS tend to be less physically active then healthy peers (13), have comparably high sedentary time as the general population (14), and commonly report sleep disturbances (15). Research in CCS focused on self-reported physical activity (16–20), which is prone to recall bias (21). Sedentary behavior has been less researched and predominantly operationalized as self-reported leisure screen-time (12, 22), neglecting other relevant domains such as non-screen based sedentary activities or sedentary time at work. Sleep duration has also been neglected, with the few existing studies relying again on self-report (23, 24).

In the general population, triaxial accelerometers are increasingly used for device-based assessment of physical activity intensities, activity types, and biological state (sleep vs. awake) (25). Such comprehensive assessment allows assessing the compositional impact of physical activity, sleep, and sedentary behavior on health outcomes instead of focusing solely on one of those behaviors (26). The few studies with device-based assessment in CCS used hip-mounted accelerometers which were only worn during waking hours or pedometers, focusing on physical activity (13, 27, 28). This leaves a critical gap regarding device-based 24-hour physical behavior assessment in CCS. With the Swiss Childhood Cancer Survivor Study (SCCSS)-Activity, we aimed to assess the feasibility of 24-hour physical behavior assessment with thigh-worn accelerometers among an age-heterogenous CCS population.

## Methods

### Study design and study population

The SCCSS-Activity is nested within the national Swiss Childhood Cancer Survivor Study (SCCSS), a population-based study of long-term outcomes among all children and adolescents registered in the Swiss Childhood Cancer Registry (ChCR). Detailed methods of the SCCSS are published elsewhere (29). The study is registered with Clinicaltrials.gov (NCT03297034). The ChCR includes all individuals diagnosed before age 20 with leukemia, lymphoma, CNS tumors, malignant solid tumors, or Langerhans cell histiocytosis in Switzerland since 1976 (30). The SCCSS repeatedly surveys survivors starting five years post-diagnosis on physical, mental, and social health, health behaviors, and quality of life, with questionnaires in German, French, and Italian; for survivors aged 5–15 years, parents respond. For SCCSS-Activity, we included survivors who participated in the 2024-2025 SCCSS mailing and were interested in participating in an accelerometer-based study. SCCSS-Activity began in February 2025 and data collection is ongoing. Ethical approval for both SCCSS and SCCSS-Activity was granted by the ethics committee of the canton of Bern (KEK-BE: 166/2014, 2021-01462, 2024-02240).

### Study procedures SCCSS-Activity

In the SCCSS questionnaire (*Figure 1, step 1*), we asked participants if they would be interested in wearing an accelerometer for one week to assess physical activity objectively (response options: yes/no). For participants responding “yes”, the SCCSS team forwarded contact details (phone, email, address) to the SCCSS study team. In case of missing responses, the SCCSS team contacted participants directly to inquire about their interest. Interested participants received study information, consent form, and a prepaid return envelope by mail consecutively depending on device availability *(Figure 1, step 2*). Earliest one week after the expected delivery of study materials, we followed up by phone or email to confirm participation. For participants for whom only a postal address was available, we requested phone/email information with the consent form and sent a reminder after two weeks if no response was received. When participants confirmed, we scheduled a measurement week that reflected their usual daily life (*Figure 1, step 3*). If the scheduled week was more than two weeks ahead, we scheduled a later call (e.g., after holidays). We mailed an accelerometer package including adhesives and replacements, illustrated mounting/dismounting instructions, a wear time protocol, a prepaid return envelope, and a return checklist (*Figure 1, step 4*). We instructed participants to mount the accelerometer immediately upon receipt (*Figure 1, step 5*), to wear it continuously for eight days, to remove it on the ninth day in the morning, and to mail it back the same day. The day after the package was due to be delivered at the participant, a study team member called to confirm mounting and to answer potential questions (*Figure 1, step 6*). After completion of the measurement period (*Figure 1, step 7*), we waited four working days to receive the accelerometer (*Figure 1, step 8*). If we had not received the device, we contacted participants by phone or email. We then downloaded the data (*Figure 1, Step 9*) and generated an individual summary report that displayed time spent physically active, sedentary and sleeping, and time spent in different activities (sleeping, sitting/lying, seated transportation, walking/running, and cycling). We sent this report together with an age-appropriate physical activity booklet (31) to the participants, and 20 CHF gift card to those who had worn the device at least 20 hours per day (*Figure 1, step 10*). To obtain feedback, we invited nine participants to a 10–15-minute semi-structured interview. We created an interview guide based on the study procedures (*Supplementary Table A.1*), asking about clarity, problems, and missing information at each step of the study. Our goal was to capture insights beyond quantitative feasibility indicators and to improve study procedures. Two researchers conducted the first interview together and compared notes. Because their notes matched, one researcher conducted the remaining eight interviews.

**Figure 1.**
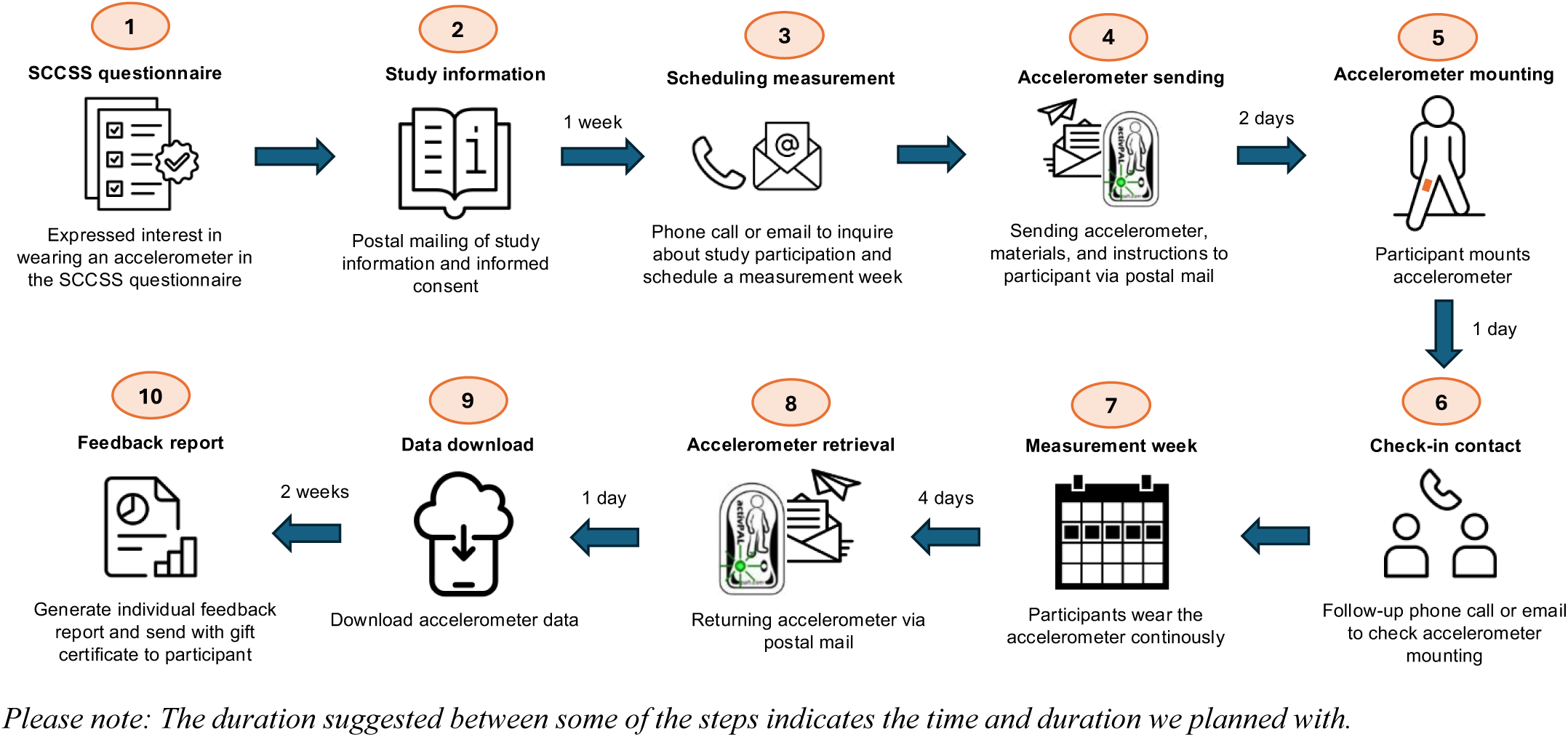
Study procedures SCCSS-Activity.

### 24-hour physical behavior measurement

We used activPAL4® devices (PAL Technologies, Glasgow, Scotland) to assess 24-hour physical behaviors. The activPAL® validly captures posture and activity type in children, adolescents, and adults (32, 33). We instructed participants to attach the device to the front of the right thigh midway between knee and hip with PALpatch® adhesives. We pre-packed each accelerometer in a nitrile sleeve and sealed it with PALpatch® materials. Participants placed an Opsite Flexifix® adhesive over the device to ensure waterproofing and prevent detachment (e.g., when changing clothes). We used the PAL Software Suite (v9.1.2.2) to download data, generate event spirals showing activity types for participant reports, and export csv-files for further processing. We defined a valid dataset as ≥20 hours of wear time on at least four weekdays and one weekend day (34, 35).

### Feasibility indicators

We chose feasibility indicators along the steps of the study procedures, including recruitment capability, acceptability and suitability of contact initiation to schedule the measurement week, accelerometer sending and returning procedures, accelerometer adherence, individual summary reports, and participant’s overall impression. *Table 1* provides a detailed overview of the feasibility categories, indicators, and data sources.

**Table 1.** Feasibility categories, data sources, indicators, and quantitative results of SCCSS-Activity.

| Feasibility category | Data sources | Indicator | Results |
| --- | --- | --- | --- |
| *Recruitment capability* | Tracking of registration, dates, and accelerometer send-out and returns | **Number of participants who completed a SCCSS baseline questionnaire in the 2024/2025 mailing** | **367** |
|  |  | N (%) of participants expressing interest in SCCSS-Activity in the SCCSS-questionnaire | 157 (43s%) |
|  |  | N (%) of participants interested in SCCSS-Activity who provided phone or email contact information in the SCCSS questionnaire | 141 (38%)<sup>a</sup> |
|  |  | N (%) of participants we invited to SCCSS-Activity | 125 (34%) |
|  |  | N (%) of participants who confirmed participation | 100 (27%) |
|  |  | N (%) of participants with scheduled measurement week | 93 (25%) |
|  |  | N (%) of participants who returned the accelerometer | 88 (24%) |
| *Acceptability and suitability of contact initiation and measurement week scheduling* | Tracking of contact efforts and dates | **Number of participants we initiated contact with:** | **125** |
|  |  | N (%) of contact type with participants after postal mailing of study information | Phone: 68 (54%)<br>Email: 28 (22%)<br>Phone + email: 29 (23%) |
|  | Interviews with participants | Median number (IQR) of phone calls and emails to reach participants | Phone calls: 2 (1-4)<br>Emails: 3 (2-4) |
|  |  | Median number of days (IQR) between the day we sent the contact information and the day we reached participants | 11 (10-19) |
|  |  | Participant's experiences regarding study information, contact channels and communication | See description in results section |
| *Suitability of accelerometer sending and returning procedures* | Tracking of dates | **Number of participants we sent accelerometers to:** | **93** |
|  |  | Median number of days (IQR) between sending and mounting the accelerometer | 1 (1-2) |
|  |  | Median number of days (IQR) between sending the accelerometer and reaching participant for follow-up contact | 3 (2-4) |
|  | Interviews with participants | Median number of days (IQR) between dismounting the accelerometer and accelerometer arrival back at the study center | 2 (2-4) |
|  |  | Number of participants we followed up due to the accelerometer not being returned as expected | 8 |
|  |  | Number of accelerometers lost or damaged | 5 |
|  |  | Participants' experiences with sensor (dis)mounting and sending procedures | See description in results section |
| *Accelerometer adherence* | Processed accelerometer data | **N (%) of participants who returned the accelerometer** | **90** |
|  |  | Median number of days (IQR) per participant that the accelerometer was worn for at least 20 hours | 7 (7-8) |
| | | N (%) of participants with valid accelerometer data ($\geq 20$ hours on at least 4 weekdays and at least 1 weekend day) | 88 (98%) |
|  | Interviews with participants | N (%) of participants returning the wear-time protocol | 89 (99%) |
|  |  | N (%) of participants filling in something in the wear-time protocol | 83 (92%) |
|  |  | Participant's experiences with wearing the accelerometer, including problems and adverse events | See description in results section |
| *Individual summary report* | Tracking of requested reports | N (%) of participants who wanted to receive a report about their individual 24-hour physical behaviors | 90 (100%) |
|  |  | N (%) of participants interested in a summary of the end study results | 89 (99%) |
|  | Interviews with participants | Participant's feedback of the individual summary reports | See description in results section |
| *Overall impression* | Interviews with participants | Participant's overall impression of the study | See description in results section |

### Statistical and qualitative analysis

We used descriptive analysis with median, interquartile range (IQR), minimum, maximum, and frequencies to assess feasibility indicators. To evaluate accelerometer adherence and wear time, we processed raw files in ActiPASS (v2025.04.2) (36) and imported the processed data into RStudio (v4.5.0) for all statistical analyses. To analyze participant interviews, we applied the framework method and created a summary based on input in the rows (37).

## Results

### Recruitment capability

*Figure 2* presents the flow diagram of the recruitment procedures. In the SCCSS 2024/2025 questionnaire, we traced and contacted 786 survivors. Of those, 367 participants returned the SCCSS questionnaire, and 157 (43%) were eligible and expressed interest in wearing an accelerometer. Interested survivors were between six and 32 years old (median: 14 years, IQR: 10-21 years); 78 (50%) were female. Of those, 104 (66%) came from the German-speaking part of Switzerland, 44 (28%) from the French-speaking, and 9 (6%) from the Italian-speaking part.

**Figure 2.**
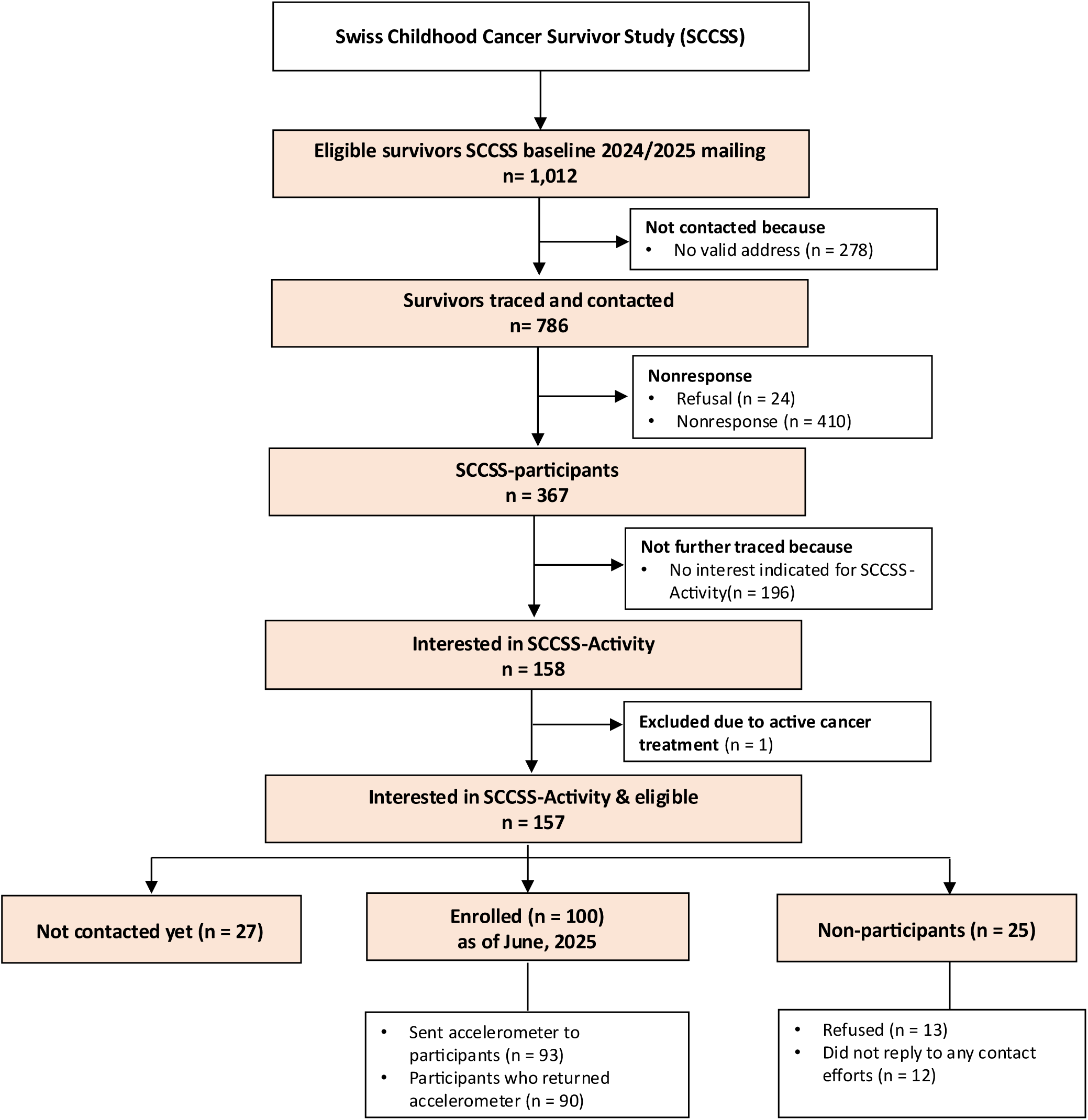
Study population tree SCCSS-Activity.

As of June 2025, we sent the study information to and followed up with 125 participants; 100 confirmed participation, 13 refused participation, and 12 did not reply to contact efforts. Of those who refused, most had physical restrictions (e.g., having to wear thigh-high compression socks), which interfered with wearing the accelerometer. We sent accelerometers to 93 participants so far; 90 returned the accelerometer (see *Table 1*). We contacted a median of five eligible participants (IQR 1-9) per week.

### Acceptability and suitability of contact initiation and measurement week scheduling

We followed up with participants earliest one week after the expected arrival of the study information, predominantly via phone. *Table 1* shows the details. In interviews, participants reported that the information was understandable and sufficient for an informed decision. They valued the one-page version for children, which enabled them to decide themselves. One participant highlighted the importance of the note that no location tracking was involved. Two participants found the information long and repetitive, leading them to read it only superficially; they recommended adding a short summary to accompany the full version. Most participants preferred phone calls to arrange participation and schedule the measurement week, citing efficiency and the chance to ask questions directly. One person preferred email, as it allowed replying at a convenient time. *Supplementary Table A.2* summarizes each interview participant’s feedback across study procedures.

### Accelerometer sending, mounting, and returning procedures

As planned, the median time between sending and mounting the accelerometer was one day (IQR 1–2), and the median retrieval time was three days (IQR 2-4) (*Table 1*). Of eight follow-ups due to delayed returns, six devices were already in transit, one was returned after a reminder, and one participant did not respond. During mailing the accelerometers back to the study center, four accelerometers were lost and one was damaged; most participants whose accelerometer got lost during volunteered for another measurement week. In interviews, participants found the illustrated instructions very helpful for mounting. Those with questions appreciated follow-up contact, while those without questions found the check-in neither useful nor disturbing. Return procedures were clear.

### Accelerometer adherence, problems, and adverse events

In interviews, most participants reported no problems wearing the accelerometer in daily life, consistent with the quantitative data: the median number of days with valid wear time duration (≥20 hours) was seven days (IQR 7–8). Eighty-eight participants (98%) provided valid datasets (≥20 hours on at least 4 weekdays and one weekend day). Some reported the need to replace adhesives when they loosened or caused slight skin irritation; minor irritations upon removal were also noted but resolved quickly.

### Individual summary report

All participants who returned the accelerometer were interested in receiving an individual summary of their 24-hour physical behaviors. In interviews, they found the summary exciting and understandable but suggested adding context by comparing results with the general population and with other survivors with similar cancer types or treatment. Several pediatric participants showed unusually high “seated transport” time as activity type, e.g., indicating that a participant was in a car or on a train while they were actually sitting in school. This was likely due to fidgeting while sitting; while it does not make a difference in overall sedentariness, parents requested a short explanation in the report. Participants also appreciated the accompanying physical activity brochure. Most parents and children reviewed it, some tried the suggestions, but most felt already physically active and did not use it further.

### Overall impression of interview participants

Interview participants described the study as positive, exciting, practical, well organized, and understandable, with minimal burden. Follow-ups made them feel comfortable and well cared for, and they valued having both email and phone as contact options. One participant noted the study reminded them that they are healthy again and able to live an active lifestyle after cancer.

## Discussion

Our study demonstrates that 24-hour physical behavior assessment with thigh-worn accelerometers is feasible among childhood cancer survivors of different age groups. Nearly half of eligible participants expressed interest in wearing the accelerometer, nearly all who wore the accelerometer returned valid data, and participants perceived study burden as minimal with high acceptability.

Nearly half (43%) of eligible CCS expressed interest in participation, similar to other population-based studies. In the German MoMo study, 46% of children and adolescents asked on-site agreed to hip-worn accelerometry (38). In the UK Biobank study, 45% responded to an email invitation for wrist-worn devices (39). The Physical Activity in Childhood Cancer Survivors (PACCS) study, which recruited CCS in clinics for hip-worn accelerometry, achieved 59% (40). Considering our mailing-based approach, our rate reflects realistic uptake among survivors.

Participants strongly accepted contact procedures, preferring phone over email. The median of two calls per participant shows that phoning survivors is feasible for confirming participation and scheduling a measurement week while allowing questions. Our accelerometer mailing procedures proved effective: Return rates were high, with only 5% of accelerometers lost or damaged, comparable to other studies (41). In-person distribution reduces loss (42), but requires greater resources when recruiting nationally. Four accelerometers broke through return envelopes during mailing, though the damaged envelopes were retrieved. We adapted mailing by using enforced padded envelopes (see *Table 2*), which should prevent future loss. Participants found illustrated instructions clear for mounting and dismounting. Although most did not view the check-in phone call as necessary, we kept it because it solved uncertainties (e.g., dismounting date) and helped address issues such as wrong addresses or delayed arrivals. When unnecessary, calls lasted less than a minute, keeping participant and staff burden minimal.

**Table 2.** Adaptations to the study procedures following the feasibility evaluation.

| Feasibility category | Original procedure | Adapted procedure |
| --- | --- | --- |
|  | In the SCCSS questionnaire, we asked the following question: | We adapted the question to provide some more information about the sensor: |
| *Recruitment capability* | We can measure physical activity more precisely with a small movement sensor. Would you be interested in wearing a movement sensor for one week as part of this study?<br>We would contact you separately for this. | **Would you be interested in having your physical activity measured with a movement sensor?**<br>We would send you a movement sensor that you attach to the thigh with a plaster. The sensor is barely noticeable and can be worn during all activities, e.g., while sleeping, showering, and exercising. Would you be interested in wearing a movement sensor for one week as part of this study? We would contact you separately about this. |
| *Acceptability and suitability of contact initiation and measurement week scheduling* | We initially contact all participants via phone. | We noticed that adolescents (14-17 years) can often better be reached via email, so we changed the preferred contact mode to emails for this age group. |
| *Suitability of accelerometer sending and returning procedures* | We sent accelerometers with so-called “B-mailing”, which is cheaper, but takes up to three days for delivery.<br>We provided participants with two envelopes to return the accelerometer: A padded small envelope for the accelerometer that was to be put in a larger envelope together with the other materials to return. | We now send accelerometer with “A-mailing”, which guarantees delivery the next day, making it easier to ensure arrival on the date agreed upon with participants.<br>Some participants did not put the accelerometer in the small padded, but only in the larger envelope. This led to some accelerometers breaking through the envelope during the postal mailing. We thus now provide participants only with a large padded return envelope, which increased returning costs, but ensures accelerometer arrival. |
| *Accelerometer adherence* | Participants attached a waterproof Opsite Flexifix® on the top of the accelerometer. | We assume that skin irritations were partially due to the waterproof, non-breathable plaster worn on top of the accelerometer. Since we exchanged our activPAL® devices against the new edition of waterproof devices, there is no nitrile sleeve or waterproof plaster needed anymore. We thus switched the waterproof plaster against a breathable tape that ensure that device does not accidentally get lost (e.g., when changing pants or during outdoor play). |
| *Individual summary report* | Participants received an individual summary report with visualizations created with PALanalysis software. | Some children had unusually high amounts of seated transport time in the event spiral. We added an explanation in the feedback report that this might be a result of the child fidgeting and does not necessarily indicate that the child was in transport. |

Accelerometer adherence was excellent: all but one participant returned valid accelerometer data. This compliance exceeds the average of 77% in accelerometer studies with children and adolescents (43). In Switzerland’s SOPHYA-study, 84% met wear time criteria (≥10 h/day on 3 weekdays and 1 weekend day) (44). In Germany’s Health Interview and Examination Survey (KIGGS), compliance reached 75% for adolescents and 78% for young adults (45).

Our higher compliance likely stems from requesting continuous 24-hour wear, which increases adherence compared to waking-time only protocols (46). Other possible reasons include the greater effort required to remove thigh-worn accelerometers compared to belt-attached hip devices, their ability to be worn during water-based activities, or participants’ pride in showing such a sportive device to others. Overall, participants reported low study burden. This aligns with studies among adult cancer survivors, who found thigh-worn devices comfortable (47). In contrast, children and adolescents have reported lack of social conformity as a burden with hip-mounted accelerometers (48). Our participants did not report this, likely because thigh-mounted devices are easier to hide under clothing. Due to minor skin irritations, we exchanged the waterproof adhesive placed on top of the accelerometer against breathable tape, probably preventing such problems in the future (see *Table 2*).

This study advances health behavior assessment with wearables by demonstrating the feasibility of 24-hour thigh-worn accelerometry among pediatric, adolescent, and adult CCS. Moving beyond physical activity, this approach enables also comprehensive assessment of device-based sedentary behavior and sleep in survivorship research (49). Limitations include potential self-selection bias, since we recruited only survivors who are already study participants of the SCCSS. We will assess differences between participants and non-participants once recruitment is complete. Because we included only recent SCCSS participants, our population was relatively young, limiting generalizability to older CCS who may find handling accelerometers less intuitive. Finally, we collected most data in spring, so we could not evaluate wearing comfort and compliance during hot temperatures.

In conclusion, thigh-worn accelerometry is a feasible and acceptable method to assess 24-hour physical behaviors among pediatric, adolescent, and adult CCS. This holds promise for future research to better understand and intervene on physical behaviors and to understand their compositional impact on CCS’ health.

## Conflict of Interest statement

CS reports a relationship to Swedish Orphan Biovitrum AB that includes travel reimbursement. This relationship has no association with the current study.

## CRediT Author Statement

CN: Conceptualization, Methodology, Formal analysis, Investigation, Visualization, Funding acquisition, Writing – original draft, Writing – review and editing

DM: Investigation, Project administration, Writing – review and editing

CS: Funding acquisition, Writing -review and editing

EB: Writing – review and editing

CK: Conceptualization, Methodology, Supervision, Funding acquisition, Writing – review and editing

## Acknowledgements

We thank all survivors for participating in our study, the study team of the Childhood Cancer Research Group, the data managers of the Swiss Paediatric Oncology Group, and the team of the Swiss Childhood Cancer Registry.

## Funding

This study was financially supported by the Swiss Cancer League and Swiss Cancer Research (KLS/KFS-482501-2019, KLS/KFS-5711-01-2022, KFS-6346-02-2025), Kinderkrebshilfe Schweiz (www.kinderkrebshilfe.ch), Kinderkrebs Schweiz (www.kinderkrebs-schweiz.ch) Stiftung für krebskranke Kinder - Regio Basiliensis (https://www.stiftung-kinderkrebs.ch). Berne University Research Foundation funded the acquisition of accelerometers.

## Data availability statement

The data that support the information of this manuscript were accessed on secured servers of the Institute of Social and Preventive Medicine at the University of Bern. Individual-level, fully anonymized, sensitive data can only be made available for researchers who fulfil the respective legal requirements. Requests of data from the Childhood Cancer Registry must be directed to the Childhood Cancer Registry of Switzerland (https://www.childhoodcancerregistry.ch). Requests of data from the Swiss Childhood Cancer Survivor Study (SCCSS) and SCCSS-Activity should be communicated to the study lead Claudia E. Kuehni.

## Abbreviations

CCS: childhood cancer survivors
ChCR: Swiss Childhood Cancer Registry
ICCC3: International Classification of Childhood Cancer, third edition
SCCSS: Swiss Childhood Cancer Survivor Study

---

**Supplementary Table A.1.** Semi-structured interview guide to inquire about participant’s perspectives regarding study procedures for SCCSS-Activity.

|  |  |
| --- | --- |
| *Introduction* | We would like to go through the course of the project step by step and discuss what can be improved at each stage and what should be kept. |
| *Optimization of study information and consent* | Think back to when you received the study information and consent form.<br>• Were the documents understandable?<br>• Was anything missing that you needed in order to make a decision? |
| *Planning contact* | About a week later, I called you to discuss whether you/your child would like to participate in the study and scheduled a measurement week.<br>• Would you have preferred to be contacted by email?<br>• Did you understand everything during the phone call?<br>• Was anything missing? |
| *Accelerometer instructions, mounting, and wearing* | Then we sent you the sensor, you/your child attached it and wore it for a week. We had another brief phone call.<br>• Were there any problems with attaching the sensor?<br>• Were the instructions understandable?<br>• Was your child / were you able to wear the sensor continuously? If not, why not?<br>• Did the sensor cause any discomfort? When and how?<br>• Was the phone call helpful? |
| *Accelerometer return procedures* | After 8 days, you/your child removed the sensor and sent it back to us.<br>• Were there any uncertainties about removing the sensor?<br>• Were there any uncertainties regarding the wear log? If it was not filled out, why not?<br>• Were the return shipping instructions clear? |
| *Individual summary report* | We then sent you an evaluation of your (your child's) physical activity behavior, along with the voucher and a small booklet with physical activity tips.<br>• Was the evaluation understandable?<br>• Was there anything else you would have liked to know? |
| *Overall impression and other feedback* | • What was your overall impression of the study?<br>• Did you expect anything different from the study?<br>• What should we keep the same?<br>• What didn't work well? (Where can we improve?)<br>• Do you have any other feedback for us? |

**Supplementary Table A.2.**
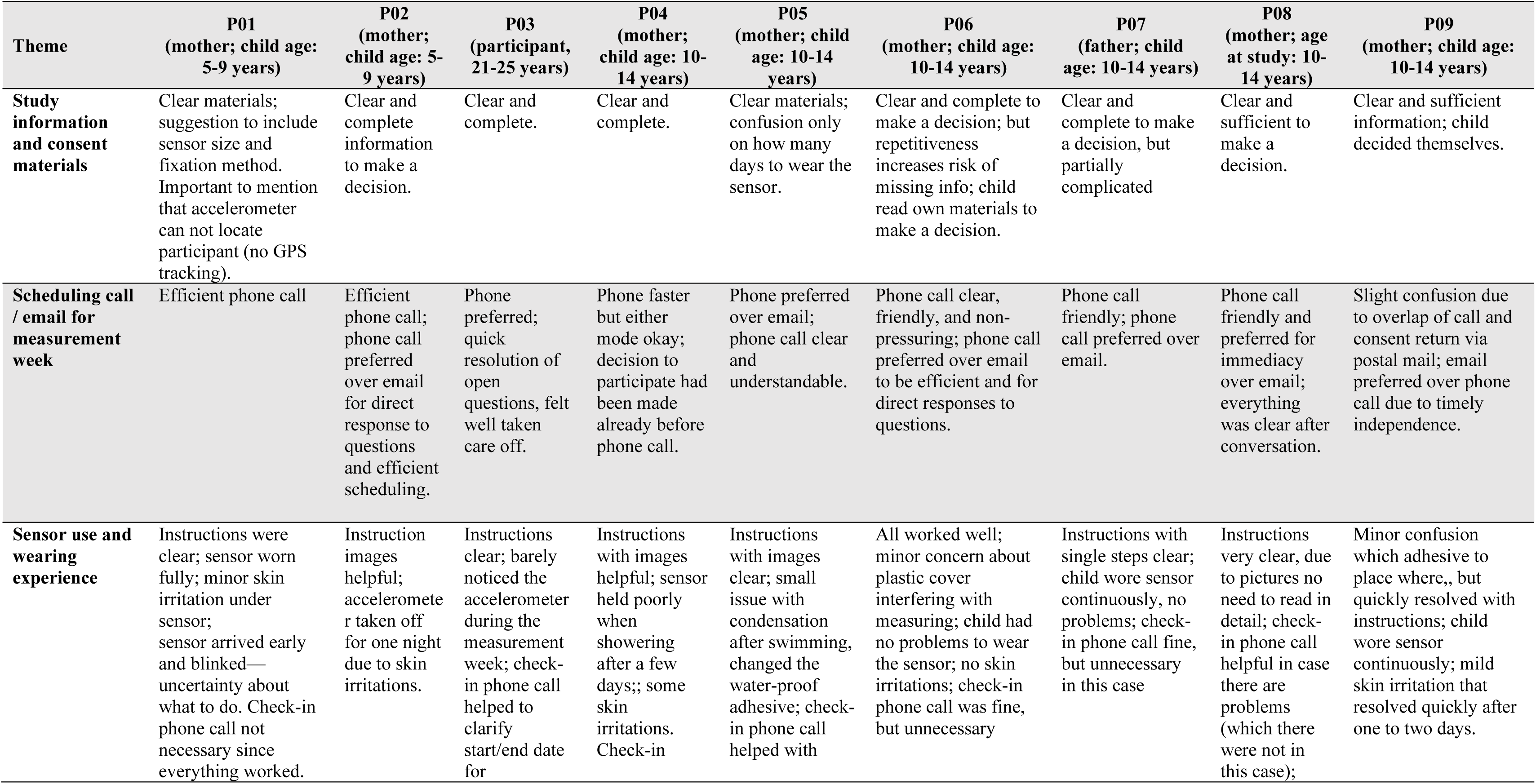
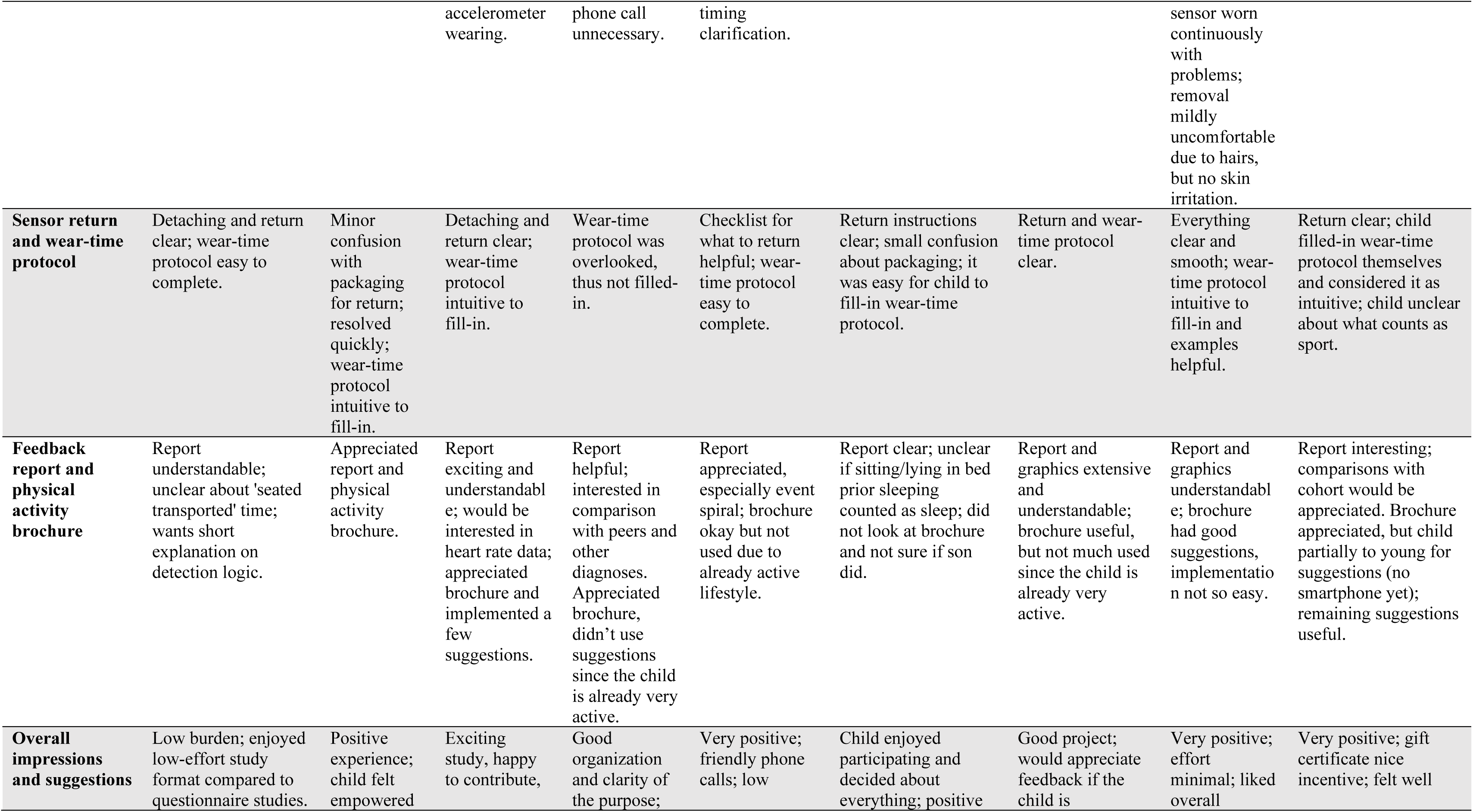
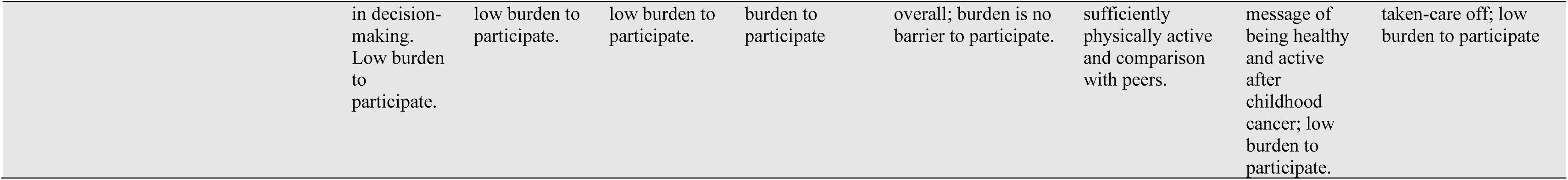
Framework matrix of participant interview summaries in SCCSS-Activity.

## Notes

### Author Declarations

Ethics committee of Canton of Bern gave ethical approval for this work.

